# Cathepsin L plays a key role in SARS-CoV-2 infection in humans and humanized mice and is a promising target for new drug development

**DOI:** 10.1101/2020.10.25.20218990

**Authors:** Miao-Miao Zhao, Wei-Li Yang, Fang-Yuan Yang, Li Zhang, Weijin Huang, Wei Hou, Changfa Fan, Ronghua Jin, Yingmei Feng, Youchun Wang, Jin-Kui Yang

**Author notes:** Correspondence (Y.F), (Y.W.), (J.-K.Y). These authors contributed equally (Miao-Miao Zhao, Wei-Li Yang, Fang-Yuan Yang, Li Zhang).

## Abstract

To discover new drugs to combat coronavirus disease 2019 (COVID-19), an understanding of the molecular basis of severe acute respiratory syndrome coronavirus 2 (SARS-CoV-2) infection is urgently needed. Here, for the first time, we report the crucial role of cathepsin L (CTSL) in patients with COVID-19. The circulating level of CTSL was elevated and was positively correlated with disease course and severity in COVID-19 patients. Correspondingly, SARS-CoV-2 pseudovirus infection increased *CTSL* expression in human cell lines and human *ACE2* transgenic mice, while *CTSL* overexpression, in turn, enhanced pseudovirus infection. CTSL functionally cleaved the SARS-CoV-2 spike protein and enhanced virus entry, as evidenced by CTSL overexpression and knockdown *in vitro* and application of CTSL inhibitor drugs *in vivo*. Furthermore, amantadine, a licensed anti-influenza drug, significantly inhibited CTSL activity and prevented SARS-CoV-2 pseudovirus infection. Therefore, CTSL is a promising target for new anti-COVID-19 drug development.

## INTRODUCTION

The recent outbreak of coronavirus disease 2019 (COVID-19) caused by severe acute respiratory syndrome coronavirus 2 (SARS-CoV-2) has imposed a severe public health crisis worldwide. To prevent and treat this disease, effective methods, such as vaccines and drugs, are urgently needed. Unfortunately, there are currently no specific therapies that can be used. Thus, the United States Food and Drug Administration (FDA) has approved remdesivir for the treatment of COVID-19 under an emergency use authorization. It is believed that remdesivir may be useful but not universally effective (Sanders et al., 2020). Therefore, broadening the spectrum of therapeutic targets is important.

Both SARS-CoV-1, which emerged in 2002, and the novel SARS-CoV-2 infect host cells through binding of their viral envelope spike (S) proteins to the same receptor, angiotensin-converting enzyme 2 (ACE2) (Hoffmann et al., 2020b). To gain entry into host cells, diverse viruses depend on cleavage and activation of the S protein by host cell proteases (Hoffmann et al., 2020a; Millet and Whittaker, 2014; Simmons et al., 2011; Sjoberg et al., 2011). Previous studies have indicated that membrane fusion of SARS-CoV-1 relies on proteolysis of the S protein by host cathepsin L (CTSL) (Huang et al., 2006; Simmons et al., 2005). Therefore, it is reasonable to hypothesize that CTSL may be a target for the development of drugs to treat SARS-CoV-2 infection (Liu et al., 2020; Smieszek et al., 2020). However, recent studies have focused primarily on furin (Hoffmann et al., 2020a) and transmembrane protease serine 2 (TMPRSS2) (Hoffmann et al., 2020b), and the effect of CTSL seems to be underestimated.

In this study, we analyzed circulating levels of CTSL in patients with COVID-19 and systematically studied the therapeutic effects of CTSL on this disease. Our data indicated that CTSL could be a promising therapeutic target for the prevention and treatment of COVID-19.

## RESULTS

### Enrollment and characteristics of patients with COVID-19

A total of 108 consecutive COVID-19 inpatients admitted to Beijing Youan Hospital, Capital Medical University between January 21 and April 30, 2020, were investigated. After excluding 5 patients who were less than 18 years old, 1 pregnant patient, 8 patients who died in a very short time, and 7 patients who were unwilling to participate, 87 patients were ultimately included in the final analysis. Of these 87 patients, 67 had nonsevere disease (2 *mild*, 65 *moderate)* and 20 had severe disease (15 *severe*, 5 *critical*) based on the severity classification established by the National Health Committee of China. Detailed demographic and clinical characteristics of the patients with COVID-19 are shown in Table S1. The marked increases in aspartate aminotransferase (AST), creatine kinase MB (CK-MB) and myoglobin levels indicated that SARS-CoV-2 attacks liver and cardiac cells, consistent with previous reports (Gupta et al., 2020; Madjid et al., 2020).

We also enrolled 125 healthy volunteers who were age- and sex-matched with the COVID-19 patients. Detailed demographic and clinical characteristics of the healthy individuals are shown in Table S2.

### The circulating level of CTSL is elevated in patients with COVID-19

As SARS-CoV-2 shares 79.6% sequence identity with SARS-CoV-1 (Zhou et al., 2020), we measured the plasma levels of several proteins that are essential to SARS-CoV-1 entry into host cells. ACE2 is the entry receptor for both SARS-CoV-1 and SARS-CoV-2. The endosomal cysteine proteases CTSL and cathepsin B (CTSB) mediate cleavage of the SARS-CoV-1 S protein, which is necessary for entry of coronavirus into host cells (Bosch et al., 2008; Simmons et al., 2005). Therefore, plasma levels of CTSL, CTSB, ACE2 and its products (angiotensin 1-7 (Ang(1-7)), were measured in patients with COVID-19. Plasma levels of Ang(1-7) were slightly lower in patients with severe disease than in patients with nonsevere disease, while plasma levels of ACE2 were the same between the two groups (Table S3). Interestingly, the plasma level of CTSL was markedly higher, while that of CTSB was slightly lower, in patients with severe disease than in patients with nonsevere disease (Figure 1A and Table S2).

**Figure 1:**
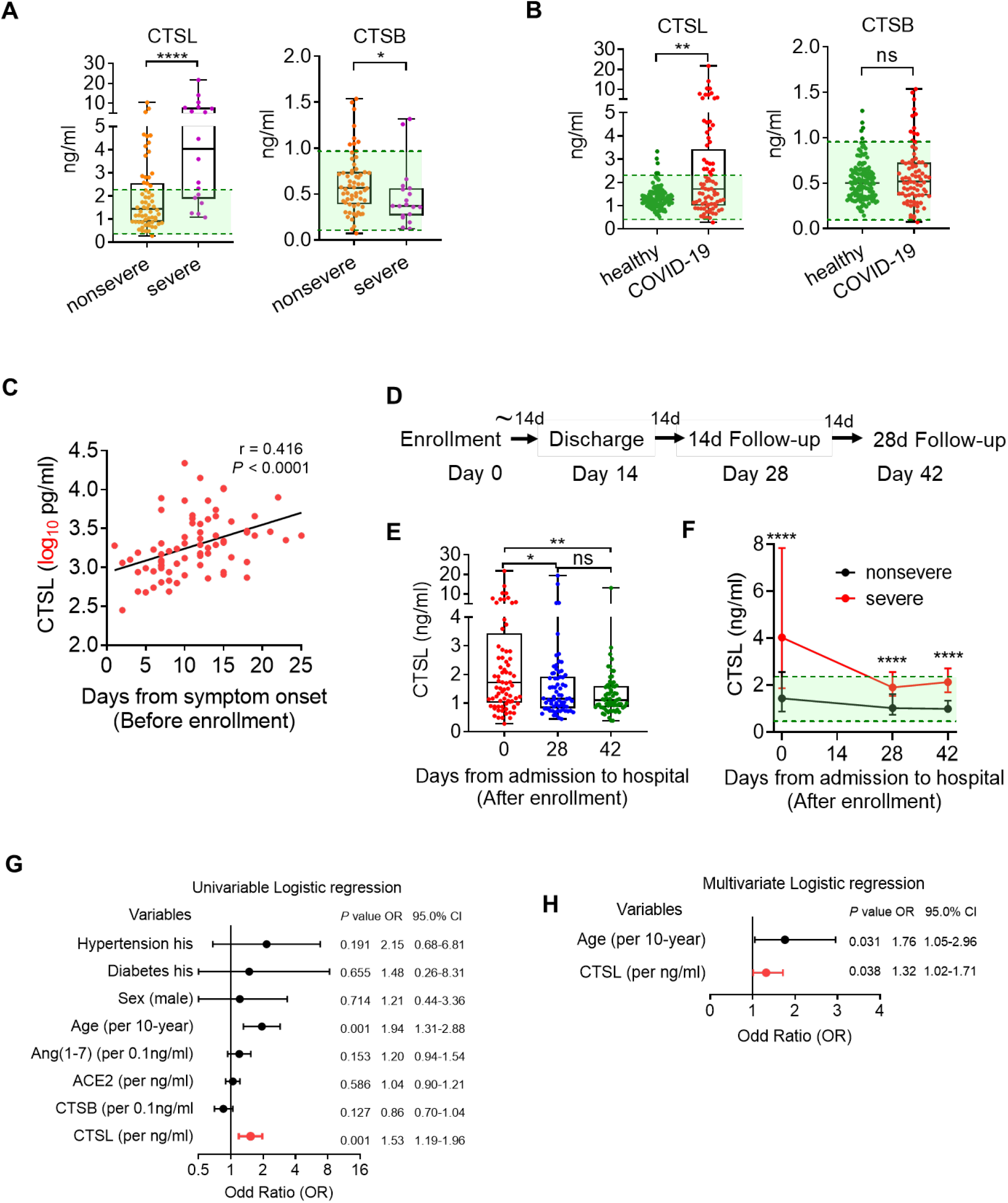
Circulating CTSL is elevated in patients with COVID-19. A total of 87 patients with COVID-19, including 20 with severe disease and 67 with nonsevere disease, and 125 healthy volunteers were enrolled in this study. **(A)** Plasma CTSL and CTSB levels patients with severe (n=20) and nonsevere COVID-19 (n=67) upon hospital admission (Day 0). Statistical significance was assessed by the Mann-Whitney *U* test (two-sided). **(B)** Plasma CTSL and CTSB levels in COVID-19 patients (n=87) and age-/sex-matched healthy volunteers (n=125). The green lines in panels A and B indicate the reference ranges for each parameter, established as the mean values ± 2 SD in the healthy participants. Statistical significance was assessed by the Mann-Whitney *U* test (two-sided). **(C)** Correlation between CTSL in plasma from COVID-19 patients (n=87) and the number of days from symptom onset to blood collection before therapy. Statistical significance was assessed by Spearman correlation analysis (two-sided). **(D)** Flowchart of the follow-up study. Patients were admitted to the hospital on Day 0 and experienced a mean hospitalization time of 14 days (Day 14). Then, they were followed up on the 14^th^ day (Day 28) and 28^th^ day (Day 42) after discharge from the hospital. Blood samples were collected on Day 0, Day 28 and Day 42. **(E)** CTSL levels in plasma from COVID-19 patients (n=87) on Day 0, Day 28 and Day 42 after enrollment. Statistical significance was assessed by the Kruskal-Wallis test with Dunn’s post hoc test. **(F)** Comparison of plasma CTSL levels between patients with nonsevere (n=67) and severe COVID-19 (n=17) on Day 0, Day 28 and Day 42. The data are shown as the medians ± interquartile ranges. Statistical significance was assessed by the Mann-Whitney *U* test (two-sided). **(G)** Summary forest plot of candidate predictor variables associated with the severity of COVID-19 (n=87) by univariable logistic regression. **(H)** Summary forest plot of candidate predictor variables associated with COVID-19 severity (n=87) in multivariable logistic regression. The predictor variables used in the final model were hypertension, diabetes, sex, age, Ang(1-7), ACE2, CTSB and CTSL.

To further confirm the correlation between changes in cathepsin levels and COVID-19, plasma CTSL and CTSB levels were measured in the 125 healthy volunteers, and the reference ranges for each parameter were established as the mean values ± 2 SD in the healthy participants, indicated by the green boxes in the figures (Figure 1A and 1B). The CTSL level was markedly higher in patients with COVID-19 than in healthy volunteers, while the CTSB level was unchanged in the patients (Figure 1B).

### The circulating level of CTSL changes with the course of COVID-19

Notably, a strong correlation was found between the CTSL level and the number of days from symptom onset to blood collection before therapy (Figure 1C). We further performed a follow-up study to determine the correlation between the CTSL level and COVID-19. Patients with COVID-19 experienced a mean hospitalization time of 14 days (Day 14) and were followed up on the 14^th^ day (Day 28) and 28^th^ day (Day 42) after hospital discharge (Figure 1D). In the follow-up study, the elevated levels of CTSL were dramatically decreased on Day 28 and remained at a stable level on Day 42 (Figure 1E). However, the difference between the severe and nonsevere groups persisted for 42 days after admission to the hospital, although in most patients, the CTSL level returned to the normal range after discharge (Figure 1F). These results indicated that CTSL was significantly associated with SARS-CoV-2 infection.

### CTSL is an independent factor for severity in patients with COVID-19

The correlations between disease severity and clinical parameters, including CTSL, CTSB, ACE2, Ang(1-7), age, sex, and coexistence of diabetes or hypertension, were estimated using Spearman’s rho correlation coefficient. Severity was positively correlated with CTSL and age but was negatively correlated with CTSB. In addition to being correlated with severity, CTSL was also positively correlated with age and history of hypertension but was negatively correlated with CTSB (Table S4). In univariable logistic regression analysis, the odds ratio (OR) of experiencing critical condition was significantly higher in patients with a higher CTSL level (OR, 1.53 per ng/ml; 95% CI, 1.19-1.96; *P* = 0.001) and older age (OR, 1.94 per 10 years; 95% CI, 1.31-2.88; *P* = 0.001), while CTSB, ACE2, Ang(1-7), sex, and coexistence of diabetes or hypertension did not significantly contribute to the odds of experiencing critical condition (Figure 1G). Multivariable logistic regression indicated that CTSL was an independent factor for severe disease status after adjustment for hypertension, diabetes, sex, age, Ang(1-7), ACE2 and CTSB (Figure 1H). Taken together, these findings led us to conclude that CTSL was highly correlated with SARS-CoV-2 infection and associated with the severity of the disease.

### The CTSL level is elevated in SARS-CoV-2 pseudovirus-infected cells *in vitro*

As the CTSL level is significantly elevated in the plasma of patients with COVID-19, we speculated that CTSL may be an important biomarker and therapeutic target for COVID-19. To test this hypothesis, we conducted a series of *in vitro* and *in vivo* experiments using a SARS-CoV-2 pseudovirus system. This pseudovirus is composed of replication-defective vesicular stomatitis virus (VSV) particles bearing SARS-CoV-2 S proteins (SARS-2-S), faithfully reflects key aspects of SARS-CoV-2 cell entry (Hoffmann et al., 2020b) as we previously reported (Nie et al., 2020) and can be used safely in biosafety level 2 (BSL-2) laboratories (Figure S1).

First, to determine the cell line with the highest susceptibility to SARS-2-S-driven entry, we compared several human cell lines, including human hepatoma cells (Huh7), human embryonic kidney cells (HEK293T), human lung adenocarcinoma cells (A549), and human lung adenocarcinoma cells (Calu-3) (Figure S2A). The luciferase assay results revealed that the Huh7 cell line had the highest susceptibility to SARS-2-S pseudovirus infection (Figure S2B-2F). In addition, the marked increases in the circulating levels of alanine aminotransferase (ALT) and AST in COVID-19 patients compared with healthy volunteers observed in our study (Table S1 and S2) and the hepatocellular injury observed in patients with COVID-19 by others suggested that SARS-CoV-2 attacks hepatocytes as target cells (Gupta et al., 2020). Therefore, Huh7 cells were selected for the subsequent experiments in this study.

Next, to verify whether CTSL expression increases after SARS-CoV-2 infection *in vitro*, the protein and mRNA levels of CTSL and CTSB were measured in Huh7 cells infected with SARS-CoV-2 pseudovirus (Figure 2A). Huh7 cells were infected with different doses of SARS-CoV-2 pseudovirus, as indicated by the luciferase activities (Figure 2B) and VSV phosphoprotein (VSV-P) mRNA levels (Figure 2C). Consistent with our clinical data, the mRNA (Figure 2D) and protein (Figure 2E) levels of CTSL increased in a dose-dependent manner after SARS-CoV-2 pseudovirus infection. These results confirmed our findings in patients with COVID-19 and indicated that SARS-CoV-2 infection caused CTSL upregulation.

**Figure 2:**
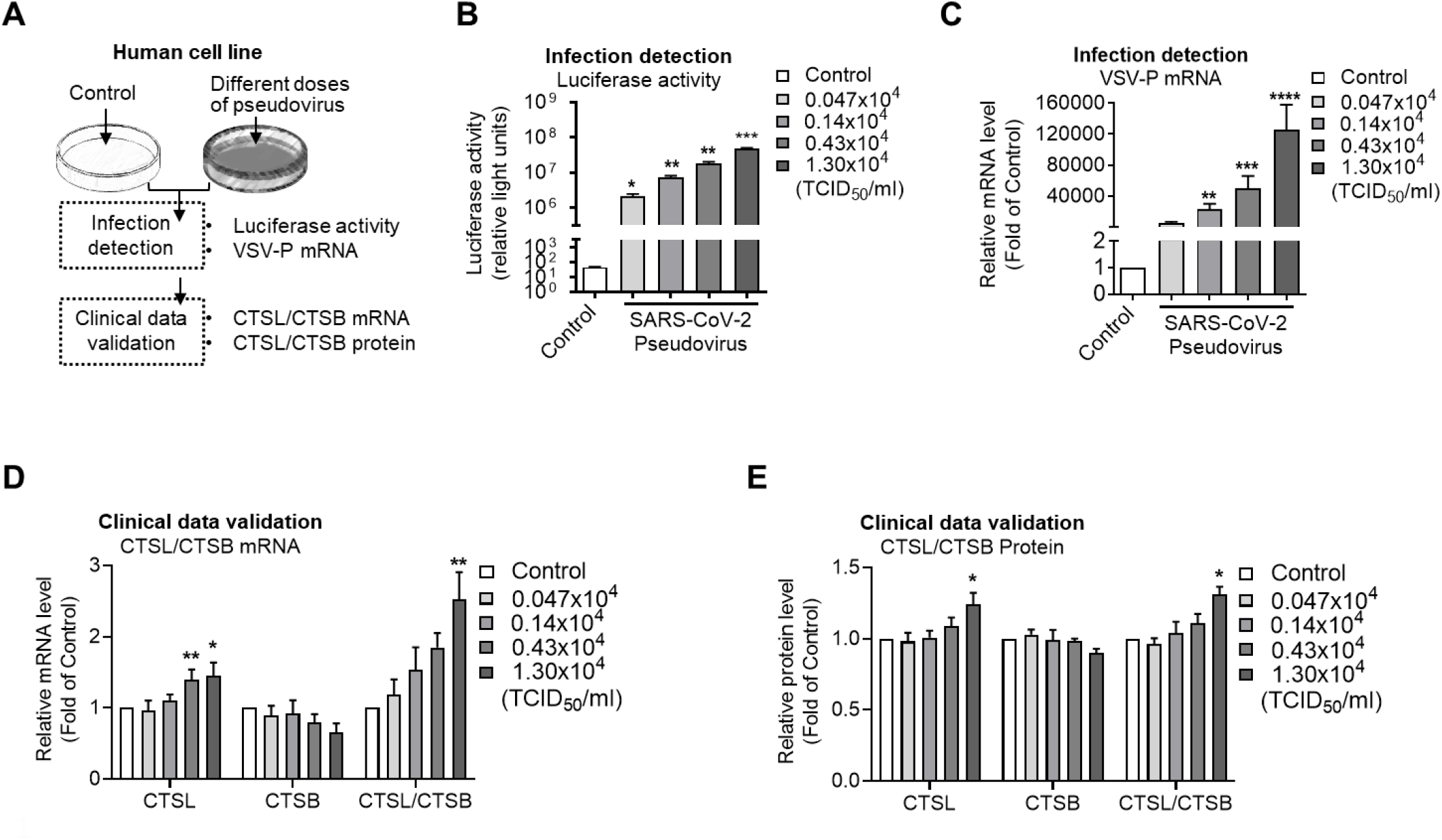
CTSL is elevated in SARS-CoV-2 pseudovirus-infected cells *in vitro*. **(A)** Schematic of the validation assay setup. Huh7 cells were infected with different doses of SARS-CoV-2 pseudovirus (from 0.047×10^4^ TCID_50_/ml to 1.30×10^4^ TCID_50_/ml). Cells not infected with pseudovirus were used as controls. Luciferase activity and VSV-P mRNA levels were measured to evaluate infection severity. The mRNA and protein levels of CTSL and CTSB in Huh7 cells were measured to validate the clinical data. **(B** and **C)** Luciferase activity (B) and VSV-P mRNA (C) levels increased dose-dependently 24 h after pseudovirus infection. n=4. Statistical significance was assessed by Brown-Forsythe and Welch’s ANOVA with Dunnett’s post hoc test in B by with the Kruskal-Wallis test with Dunn’s post hoc test for C. **(D** and **E)** Effects of SARS-CoV-2 pseudovirus infection on CTSL and CTSB mRNA levels (D) and protein levels (E). n=4-6. Statistical significance was assessed by the Kruskal-Wallis test with Dunn’s post hoc test. Data are expressed as the mean ± s.e.m. values. \**P*<0.05, \*\**P* < 0.01, \*\*\**P*<0.001, \*\*\*\**P*<0.0001.

### CTSL knockdown or overexpression affects pseudovirus infection *in vitro*

To investigate whether CTSL is required for cell entry of SARS-CoV-2, we used siRNAs against human *CTSL* (si-CTSL) and plasmids encoding human *CTSL* (pCTSL) to knockdown and overexpress the *CTSL* gene in Huh7 cells, respectively, as shown in Figure 3A. si-CTSL treatment dose-dependently downregulated CTSL without affecting *CTSB* expression (Figure 3B). Knockdown of *CTSL* led to a significant dose-dependent reduction in pseudovirus cell entry, as evidenced by the luciferase activity and VSV-P mRNA level (Figure 3C-3E). In contrast, overexpression of *CTSL* markedly increased pseudovirus cell entry in a dose-dependent manner without affecting *CTSB* expression (Figure 3F-3I). These results suggested that CTSL but not CTSB was critical for SARS-CoV-2 infection.

**Figure 3:**
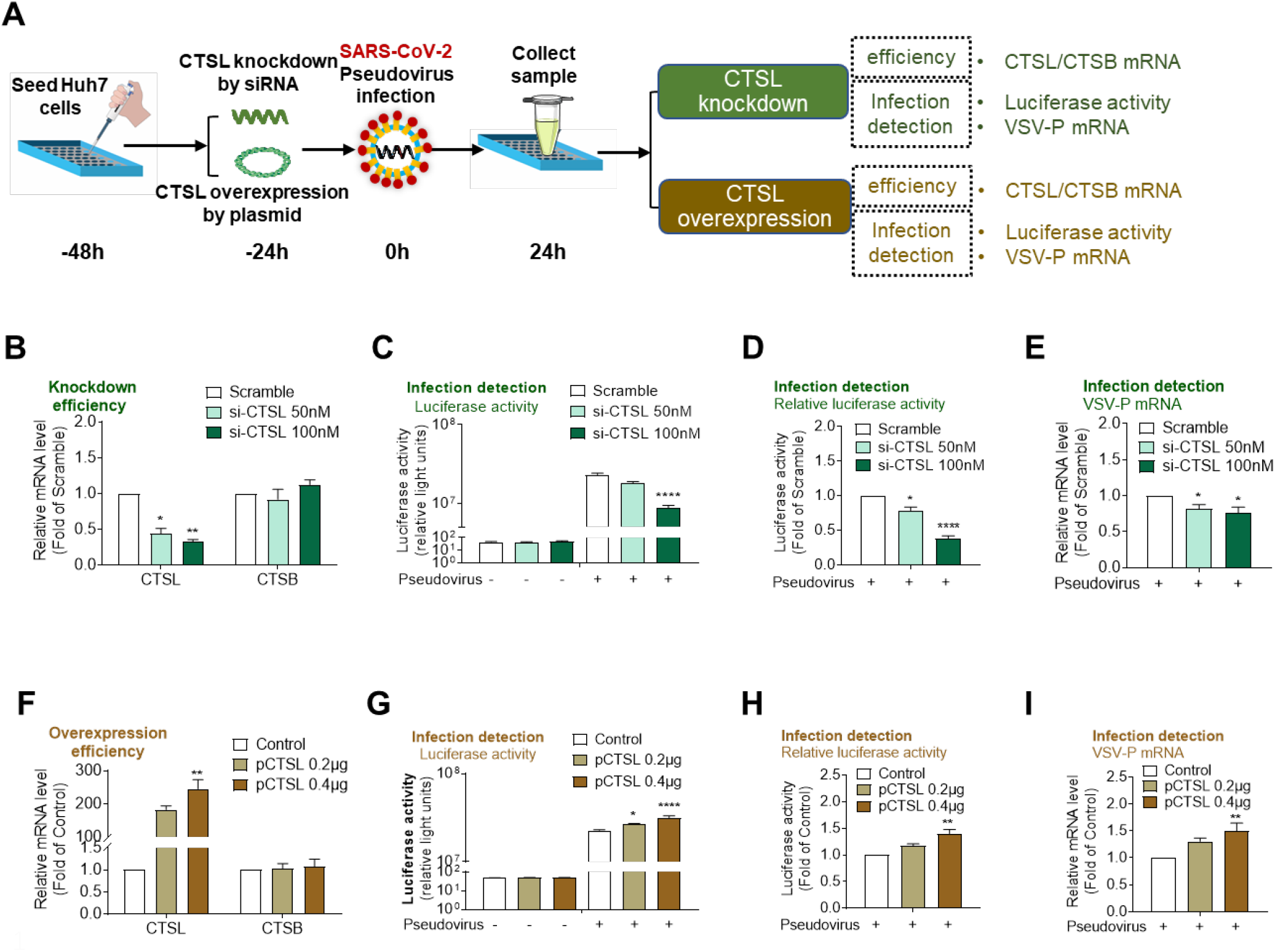
CTSL knockdown or overexpression affects pseudovirus infection *in vitro*. **(A)** Schematic of the CTSL knockdown and overexpression assay setup. **(B)** Dose-dependent knockdown of *CTSL* by siRNAs without affecting *CTSB* expression. n=4. Statistical significance was assessed by the Kruskal-Wallis test with Dunn’s post hoc test. **(C-E)** Knockdown of *CTSL* dose-dependently inhibited SARS-2-S-driven entry, as measured by a luciferase assay and shown as absolute luciferase activity (C) and relative luciferase activity (D) values, and VSV-P mRNA levels (E). n=6-8. Statistical significance was assessed by the Kruskal-Wallis test with Dunn’s post hoc test. **(F)** Dose-dependent overexpression of *CTSL* with a plasmid encoding the *CTSL* gene without affecting *CTSB* expression. n=5. Statistical significance was assessed by the Kruskal-Wallis test with Dunn’s post hoc test. **(G-I)** Overexpression of *CTSL* dose-dependently promoted SARS-2-S-driven entry, as measured by a luciferase assay and shown as absolute luciferase activity (G) and relative luciferase activity **(H)** values, and VSV-P mRNA levels (I). n=5. Statistical significance was assessed by the Kruskal-Wallis test with Dunn’s post hoc test. The data are expressed as the mean ± s.e.m. values. \**P*<0.05, \*\**P* < 0.01, \*\*\**P*<0.001, \*\*\*\**P*<0.0001.

### CTSL cleaves the S protein, and this cleavage promotes cell-cell fusion

CTSL cleaves the SARS-CoV-1 S protein into S1 and S2 subunits and proteolytically activates cell-cell fusion (Simmons et al., 2005). Unlike the SARS-CoV-1 S protein, the SARS-CoV-2 S protein is precleaved by the proprotein convertase furin at the S1/S2 cleavage site (Figure 4A). Hence, the effects of CTSL in SARS-CoV-2 seem to be replaced by those of furin. However, no report has examined the effects of CTSL on SARS-CoV-2 S protein cleavage and the function of CTSL in cell-cell fusion. Here, we directly detected the cleavage of purified SARS-CoV-2 S protein by CTSL. Treatment with CTSL resulted in successful cleavage of purified SARS-CoV-1 S protein, suggesting that the experimental system was feasible. Notably, CTSL also efficiently cleaved purified SARS-CoV-2 S protein in a dose-dependent manner (Figure 4B). To further confirm the specificity, CTSL inhibitors were employed. Given that no currently available drug specifically inhibits CTSL (Dana and Pathak, 2020), two compounds that have been demonstrated to have inhibitory activity against CTSL (E64d, a broad-spectrum cathepsin inhibitor, and SID 26681509, a relatively selective CTSL inhibitor) were used. The cleavage activity of CTSL was blocked by E64d and SID 26681509 (Figure 4B). These results indicated that CTSL efficiently cleaved the SARS-CoV-2 S protein into smaller fragments after its initial cleavage by furin.

**Figure 4:**
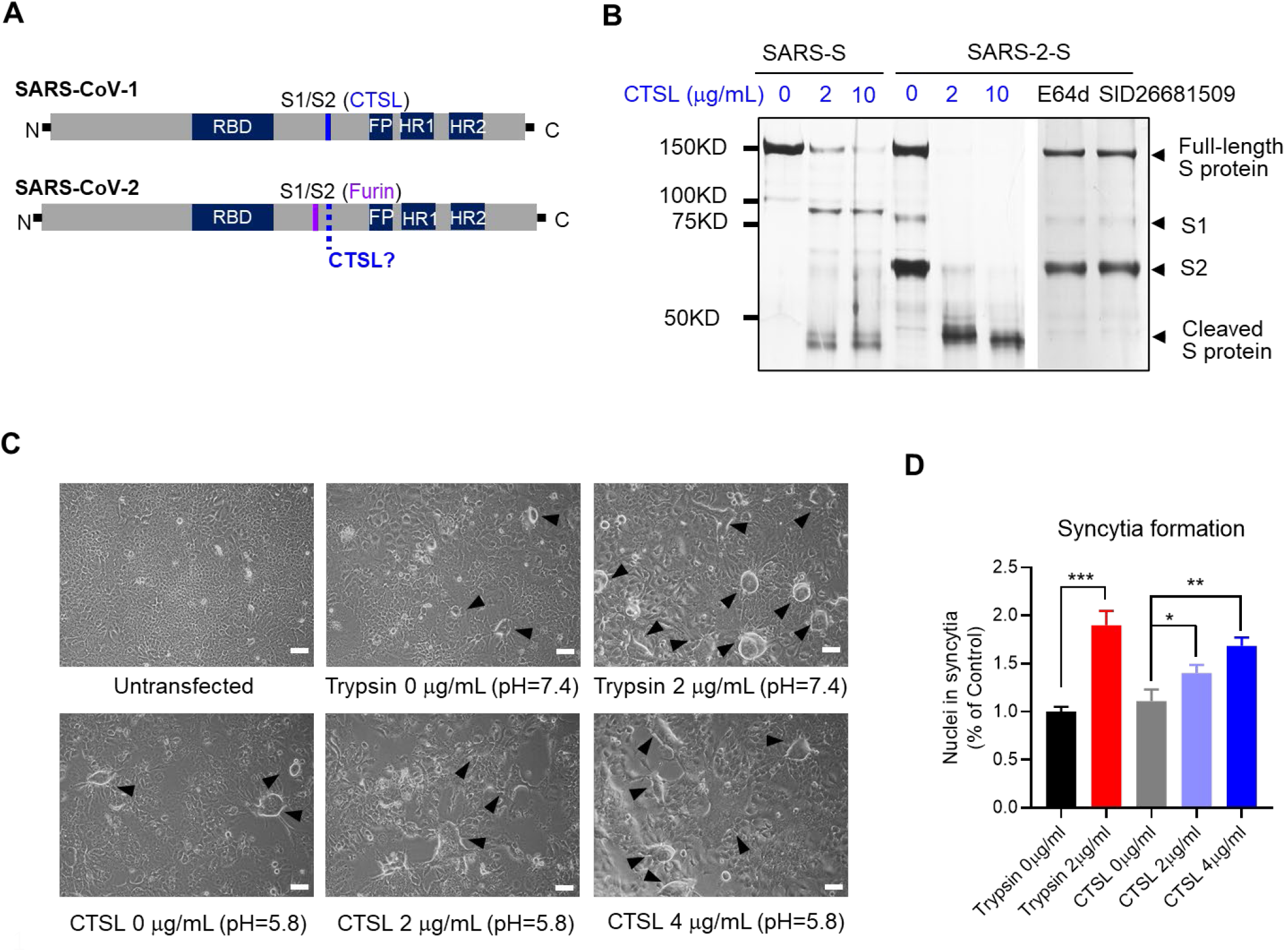
CTSL cleaves the SARS-CoV-2 spike (S) protein, and this cleavage promotes cell-cell fusion. **(A)** Overview of the SARS-CoV-1 and SARS-CoV-2 S1/S2 cleavage sites. FP (fusion peptide), HR1 (heptad repeat 1) and HR2 (heptad repeat 2) are units of the S2 subunit that function in membrane fusion. **(B)** Analysis of CTSL-mediated S protein cleavage. Purified SARS-CoV-1 or SARS-CoV-2 S protein was incubated in the presence or absence (assay buffer, pH=5.5) of CTSL (2 or 10 μg/ml in assay buffer, pH=5.5) at 37°C for 1 h. The reaction system of 2 μg/ml CTSL was further supplemented with CTSL inhibitors (20 μM E64d or 20 μM SID 26681509) as indicated. Proteins were subjected to SDS-PAGE and detected by silver staining. Representative data from three independent experiments are shown. **(C)** Syncytium formation assay: Huh7 cells were untransfected (Null) or transfected with plasmid to express the SARS-CoV-2 S protein. Cells were incubated in the presence or absence (PBS, pH=7.4) of trypsin (2 μg/ml in PBS, pH=7.4) or in the presence or absence (PBS, pH=5.8) of CTSL (2 or 4 μg/ml in PBS, pH=5.8) for 20 min. Images were acquired after an additional 16 h incubation in medium. (scale bars, 50 μm). The black arrowheads indicate syncytia. Representative data from seven independent experiments are shown. **(D)** Quantitative analysis of syncytia in panel C. n=7. Statistical significance was assessed by one-way ANOVA with Tukey’s post hoc test. The data are expressed as the mean ± s.e.m. values. \**P*<0.05, \*\**P* < 0.01, \*\*\**P*<0.001.

To investigate whether CTSL functionally cleaves the SARS-CoV-2 S protein, we performed a cell-cell fusion assay by recording SARS-2 S protein-driven formation of multinucleated giant cells (syncytium). No syncytia were observed without S protein expression, while SARS-2-S expression alone (treated with PBS, pH=7.4) resulted in syncytium formation. Trypsin treatment (2 μg/ml in PBS, pH=7.4), which has been shown to induce S-protein-mediated cell-cell fusion (Bosch et al., 2008; Ou et al., 2020), markedly increased syncytium formation nearly two-fold. In addition, compared to mock treatment (PBS, pH=5.8), CTSL (in PBS, pH=5.8) dose-dependently induced an increase in syncytium formation of up to ∼70%. These data indicated that CTSL activity, not acidic conditions, was responsible for the increase in syncytium formation (Figure 4C and 4D). Therefore, these results led us to conclude that CTSL efficiently cleaved the SARS-CoV-2 S protein and that this cleavage promoted S-protein-mediated cell-cell fusion.

### CTSL inhibitors prevent SARS-CoV-2 pseudovirus infection *in vitro*

To further confirm the role of CTSL in SARS-CoV-2 infection, Huh7 cells were treated with CTSL inhibitors, as shown in Figure 5A. Both SID 26681509 and E64d significantly inhibited SARS-CoV-2 pseudovirus infection. As E64d exhibited less cytotoxicity than SID 26681509, it was selected for the subsequent experiments (Figure 5B and 5C).

**Figure 5:**
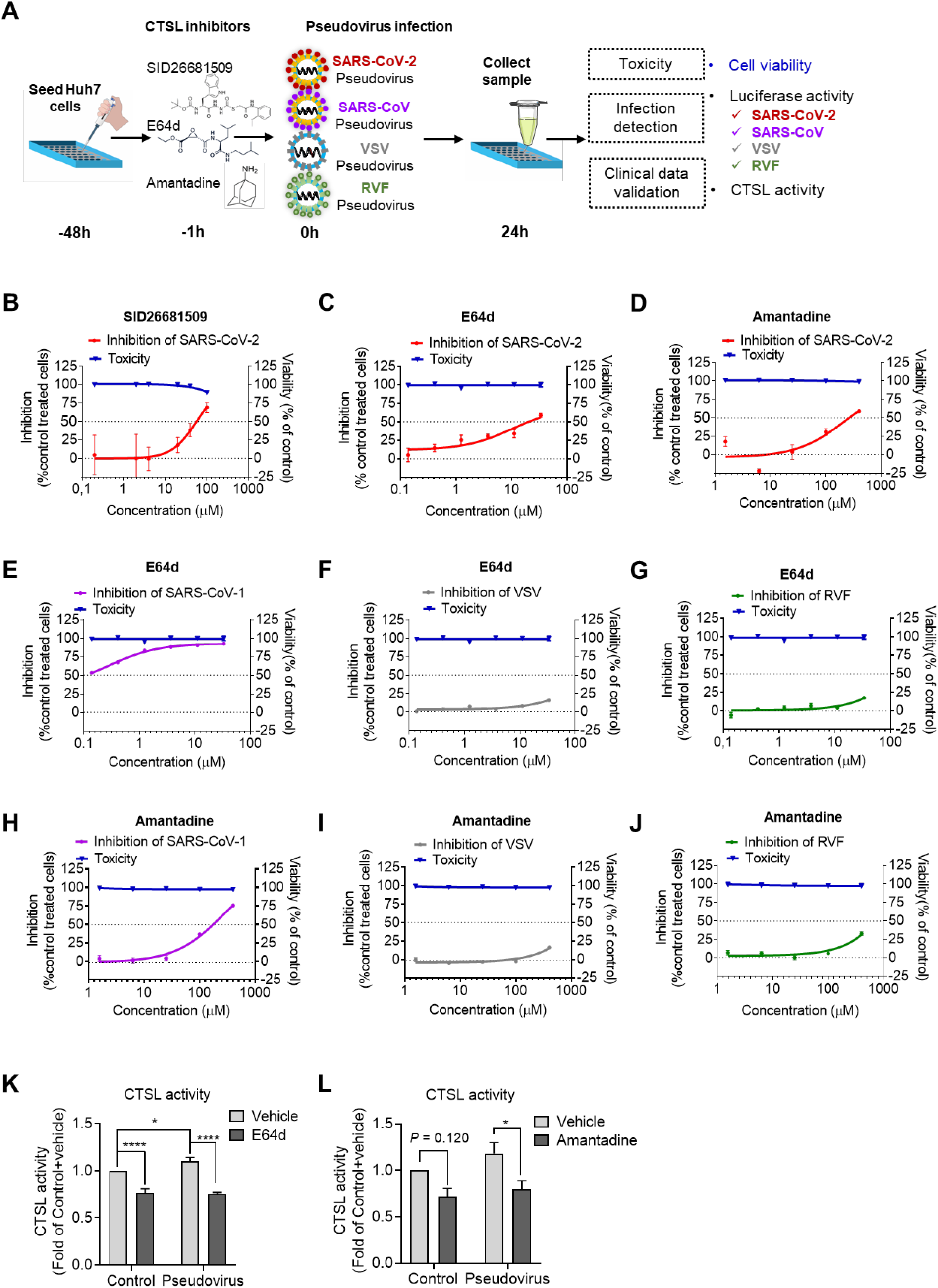
CTSL inhibitors block SARS-2-S-driven cell entry *in vitro*. **(A)** Schematic of the CTSL inhibitor assay setup. Huh7 cells were pretreated with different drugs 1 h before infection with different pseudoviruses, as indicated, at the same dose (1.3×10^4^ TCID_50_/ml). Pseudovirus infection and cell viability were evaluated by a luciferase activity and MTT assay, respectively. **(B-D)** Inhibition of SARS-2-S-driven cell entry by different doses of SID26681509 (B), E64d (C) and amantadine (D) and viability of cells treated with different doses of the drugs as indicated. n=4. **(E-G)** Effects of E64d on SARS-CoV-1 pseudovirus (E), VSV pseudovirus (F) and RVF pseudovirus (G) infection and viability of cells treated with different doses of E64d as indicated. n=4. **(H-J)** Effects of amantadine on SARS-CoV-1 pseudovirus (H), VSV pseudovirus (I) and RVF pseudovirus (J) infection and viability of cells treated with different doses of amantadine as indicated. n=4. **(K** and **L)** Effects of drug pretreatment on CTSL enzyme activity in Huh7 cells with or without pseudovirus infection. Huh7 cells were pretreated with vehicle, 30 μM E64d (K) or 300 μM amantadine (L) for 1 h and were then infected with SARS-CoV-2 pseudovirus at a dose of 1.3×10^4^ TCID_50_/ml. Cells not infected with pseudovirus were used as controls. n=4. Statistical significance was assessed by two-way ANOVA with the Holm-Sidak post hoc test for multiple comparisons. The data are expressed as the mean ± s.e.m. values. \**P*<0.05, \*\**P* < 0.01, \*\*\**P*<0.001, \*\*\*\**P*<0.0001.

Furthermore, we were interested to find that amantadine, a prophylactic agent approved by the US FDA in 1968 for influenza and later for Parkinson’s disease, has been reported to suppress the gene transcription of *CTSL* (Smieszek et al., 2020). We next examined the impact of amantadine on SARS-CoV-2 infection and found that it significantly inhibited pseudovirus infection with little cytotoxicity (Figure 5D). Moreover, both E64d and amantadine also significantly prevented SARS-CoV-1 S protein-driven but not VSV G protein-driven or Rift Valley fever (RVF) virus (a member of the bunyavirus family that does not require CTSL for cell entry (Hofmann et al., 2013; Spiegel et al., 2016)) G protein-driven pseudovirus infection (Figure 5E-5J).

Finally, CTSL enzyme activity was measured in of Huh7 cells. Treatment with E64d (Figure 5K) and amantadine (Figure 5L) inhibited the enzyme activity and blocked the activation of CTSL induced by SARS-CoV-2 pseudovirus infection. These results indicated that the therapeutic effects of both E64d and amantadine were at least partially mediated by inhibition of CTSL enzyme activity.

### CTSL inhibitors prevent pseudovirus infection in humanized mice

To further verify whether CTSL inhibition can prevent pseudovirus infection *in vivo*, the effects of E64d and amantadine on preventing SARS-CoV-2 pseudovirus infection were assessed in mice by bioluminescence imaging (BLI). Because SARS-CoV-2 recognizes the human ACE2 protein but not mouse or rat ACE2 as the cell entry receptor (Wan et al., 2020), *hACE2* humanized mice—model mice engineered to express *hACE2* via CRISPR/Cas9 knock-in technology, as we previously reported (Sun et al., 2020)—were employed. The *hACE2* humanized mice were randomly divided into four groups and treated with either vehicle or different drugs as indicated. Bioluminescence was measured and visualized in pseudocolor as an indicator of SARS-CoV-2 pseudovirus infection severity. Pseudovirus-infected humanized mice showed a significantly higher luminescence signal than healthy control mice, indicating that the mice were successfully infected (Figure 6A and 6B). Compared to the vehicle treatment, E64d significantly prevented SARS-CoV-2 pseudovirus infection. Amantadine also showed suppressive effects on pseudovirus infection, but the differences were not statistically significant (*P* = 0.058) (Figure 6B). The hepatic VSV-P mRNA level was markedly increased in humanized mice after SARS-CoV-2 pseudovirus infection, but this increase was significantly suppressed by pretreatment with either E64d or amantadine (Figure 6C), indicating that both drugs indeed prevented SARS-CoV-2 pseudovirus infection.

**Figure 6:**
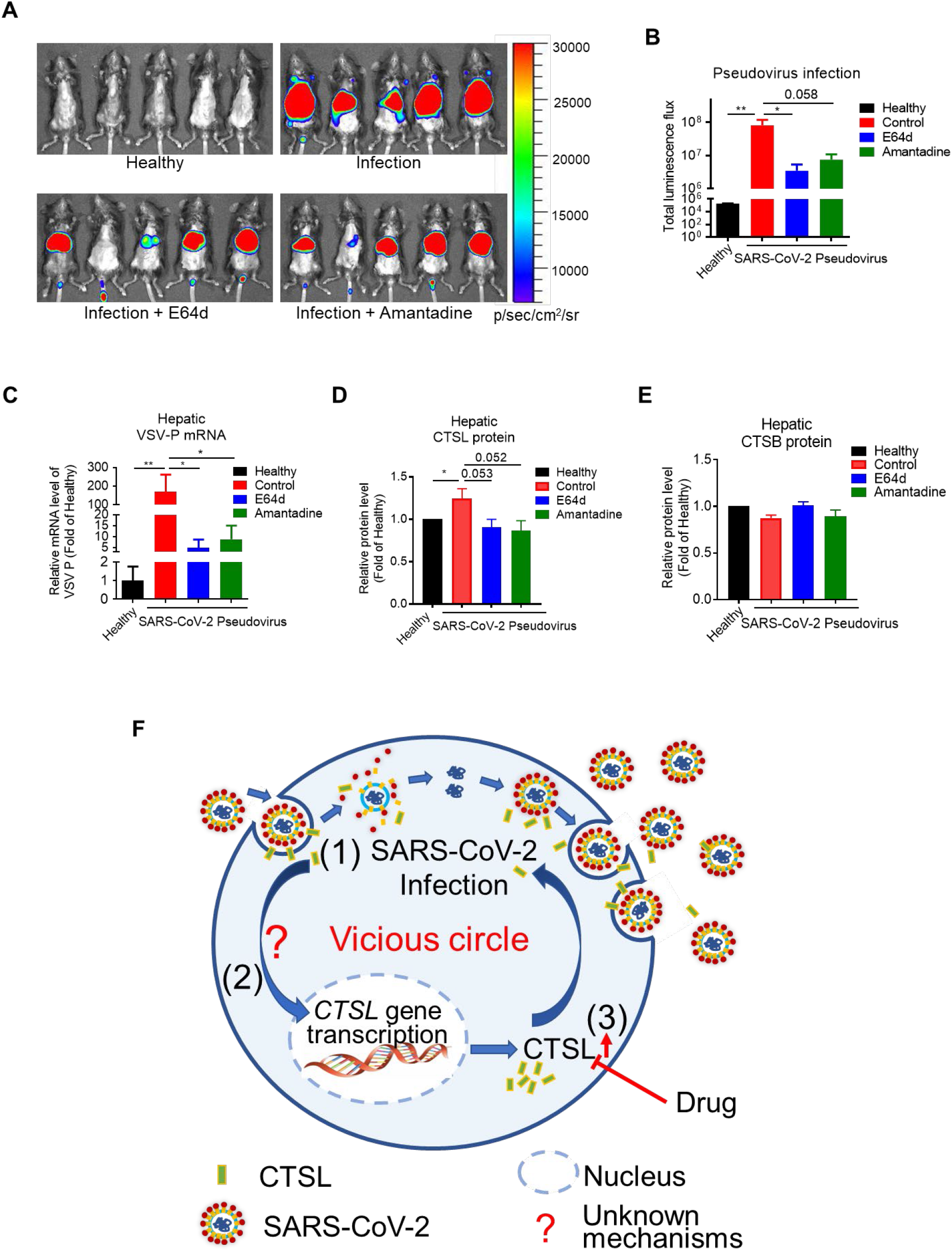
CTSL inhibitors prevent pseudovirus infection in humanized mice. Human *ACE2* transgenic mice were randomly divided into four groups and pretreated with vehicle or different drugs (E64d or amantadine) as indicated 2 days prior to virus inoculation via tail vein injection (1.5×10^6^ TCID_50_ per mouse). Mice without pseudovirus inoculation were used as the healthy control group. Bioluminescence was measured one day post infection and visualized in pseudocolor. **(A)** The relative intensities of emitted light are presented as the photon flux values in photon/(sec/cm^2^/sr) and displayed as pseudocolor images, with colors ranging from blue (lowest intensity) to red (highest intensity). **(B)** Pseudovirus infection in each group as indicated by the total flux values. Statistical significance was assessed by one-way ANOVA with Tukey’s post hoc test for multiple comparisons. **(C)** Pseudovirus infection as indicated by the liver VSV-P mRNA levels in each group. Statistical significance was assessed by one-way ANOVA with Tukey’s post hoc test for multiple comparisons. **(D)** Hepatic CTSL protein levels in each group. Statistical significance was assessed by the Kruskal-Wallis test with Dunn’s post hoc test. **(E)** Hepatic CTSB protein levels in each group. Statistical significance was assessed by the Kruskal-Wallis test with Dunn’s post hoc test. **(F)** Proposed mechanism of CTSL action in SARS-CoV-2 infection. (1) CTSL cleaves the SARS-2-S protein and releases the virus from the endosome. (2) SARS-CoV-2 promotes CTSL gene transcription and enzyme activity through unknown mechanisms. (3) Upregulation of CTSL, in turn, enhances SARS-CoV-2 infection. n=5. The data are expressed as the mean ± s.e.m. values. \**P*<0.05, \*\**P* < 0.01.

Notably, the protein level of CTSL in the liver was significantly increased in SARS-CoV-2-infected mice; this increase was reversed by treatment with either E64d or amantadine (Figure 6D), while the CTSB level was not significantly affected (Figure 6E).

## DISCUSSION

Diverse viruses, including SARS-CoV-1, have been shown to affect the expression of host cell infection-related genes (Dinan et al., 2019; Peng et al., 2010; Zhang et al., 2013). Here, for the first time, we found that SARS-CoV-2 infection promoted the gene expression of *CTSL* both *in vivo* and *in vitro*, while *CTSL* overexpression, in turn, enhanced pseudovirus infection in human cells. The detailed mechanisms of this “vicious circle” remain to be investigated. Interestingly, a recent study also found that SARS-CoV-2 can exploit interferon-driven upregulation of ACE2 to enhance infection (Ziegler et al., 2020), suggesting that the mechanistic involvement of other infection-related genes in SARS-CoV-2 infection requires further exploration. However, CTSL would be a promising therapeutic target for inhibitors that could not only inhibit entry of the virus but also block the vicious circle (Figure 6F).

Infection of cells with many kinds of viruses depends on specific host cell proteases (Reinke et al., 2017; Sakai et al., 2014). A recent study suggested that CTSL might be involved in SARS-CoV-2 entry into HEK293 cells *in vitro* (Ou et al., 2020). However, clinical evidence of the role of CTSL in SARS-CoV-2 infection is lacking. First, no investigation of circulating levels of CTSL in patients with COVID-19 had been reported before this study. Second, tissue expression of CTSL has not yet been investigated in SARS-CoV-2 infection, although circulating levels of CTSL can reflect its expression profile in many organ tissues, such as vascular tissues (Liu et al., 2006). In this study, we first analyzed the circulating level of CTSL in patients with COVID-19 and found that it precisely reflected the severity and status of COVID-19. This relationship may be attributed to the elevated expression of CTSL observed in pseudovirus-infected human cells and mouse livers.

Previous studies have suggested that CTSL inhibitors effectively prevent the infection of many other coronaviruses, including SARS-CoV-1, Middle East respiratory syndrome coronavirus (MERS-CoV) and human coronavirus (HCoV)-229E (Bertram et al., 2013; Shah et al., 2010; Zhou et al., 2016). CTSL is thought to be a potential target for the treatment of COVID-19 (Liu et al., 2020; Smieszek et al., 2020), although no systematic study has been conducted. Previous *in vitro* studies have indicated that TMPRSS2 is essential to SARS-CoV-2 infection (Hoffmann et al., 2020b). In addition, the SARS-CoV-2 S protein is precleaved by furin at the S1/S2 site, which likely reduces its dependence on CTSL (Hoffmann et al., 2020a). In this study, we showed that CTSL functionally cleaved the SARS-CoV-2 S protein into smaller fragments. This cleavage resulted in an up to ∼70% enhancement in S-protein-mediated cell-cell fusion. CTSL efficiently enhanced SARS-CoV-2 infection, as evidenced by our overexpression and knockdown data *in vitro* and inhibition data *in vivo*. Therefore, it is reasonable to conclude that CTSL, TMPRSS2 and furin are all required for SARS-CoV-2 infection. Broadening the range of therapeutic targets for COVID-19 is important, and the effects of CTSL should not be underestimated.

The COVID-19 pandemic has motivated the most immense efforts to date to identify drugs that can safely, quickly, and effectively reduce morbidity and mortality. Focusing on repurposing of a licensed drug for COVID-19 may be more efficient than starting with a preclinical drug. Thus, repurposing of many FDA-approved drugs for COVID-19 has been suggested, e.g., as was accomplished with the antimalarial drugs chloroquine (CQ) and hydroxychloroquine (HCQ) for rheumatoid arthritis and with the anti-influenza drug amantadine for Parkinson’s disease (Ballout et al., 2020; Gautret et al., 2020; Mitja and Clotet, 2020).

Amantadine is a preventive agent first used for influenza and later for Parkinson’s disease. Amantadine has been hypothesized to have therapeutic utility for COVID-19 partially because of its ability to downregulate CTSL (Aranda Abreu et al., 2020; Araujo et al., 2020; Smieszek et al., 2020). In this study, we systematically investigated the role of amantadine in the treatment of COVID-19 and found that amantadine inhibited CTSL enzyme activity in the setting of SARS-CoV-2 pseudovirus infection. Amantadine significantly inhibited SARS-CoV-1 and SARS-CoV-2 cell entry with little cytotoxicity. Moreover, amantadine showed anti-pseudovirus effects, with downregulation of CTSL expression, in humanized mice. Therefore, amantadine may be a potent therapeutic drug.

In conclusion, we report that SARS-CoV-2 infection promoted CTSL expression and enzyme activity, which, in turn, enhanced viral infection. CTSL functionally cleaved the SARS-CoV-2 S protein and enhanced viral entry. Hence, CTSL is likely an important therapeutic target for COVID-19. Furthermore, we showed that amantadine, a licensed anti-influenza drug, significantly inhibited CTSL activity after SARS-CoV-2 pseudovirus infection and prevented infection both *in vitro* and *in vivo*. Therefore, clinical trial of amantadine or other CTSL inhibitors for COVID-19 treatment would be a worthwhile endeavor.

## STAR*METHODS

### Detailed methods are provided in the online version of this paper and include the following

- KEY RESOURCES TABLE
- RESOURCE AVAILABILITY
  ○ Lead Contact
  ○ Materials Availability
  ○ Data and Code Availability
- EXPERIMENTAL MODELS AND SUBJECT DETAILS
  ○ Participants and clinical samples
  ○ Cell lines and reagents
  ○ Experimental Mice
- METHOD DETAILS
  ○ Serum biomarkers of patients with COVID-19
  ○ Pseudovirus
  ○ Luciferase assay
  ○ Cell line selection and clinical data verification *in vitro*
  ○ Cleavage of SARS-CoV-2 S protein by CTSL
  ○ Syncytium formation assay
  ○ CTSL knockdown by siRNA and overexpression by plasmid *in vitro*
  ○ Effect of drug treatment on SARS-CoV-2 entry *in vitro* and cell viability assay
  ○ Animal experiments
  ○ RNA extraction and Real-time PCR
- QUANTIFICATION AND STATISTICAL ANALYSIS

## Data Availability

The data that support the findings of this study are available from the corresponding author upon reasonable request.

## SUPPLEMENTAL INFORMATION

STAR Methods Figures S1 and S2

Tables S1 to S4

## Author contributions

J.-K.Y. conceived the idea for the study, supported the study, designed the experiments, and wrote the manuscript. M.-M.Z. W.-L.Y designed, performed the experiments, and wrote the first version of manuscript. F.-Y.Y. and L.Z. designed and performed the experiments. W.-J.H., W.H., C.-F.F., R.-H.J. partially performed the experiments. Y.-M.F, Y.-C.W. helped with the interpretation of the results, design of experiments, and proof of the manuscript.

## Acknowledgments

This work was supported by grants from National Key R&D Program of China (2017YFC0909600) and National Natural Science Foundation of China (8151101058, 81930019, 81471014) to J.K.Y.

## Ethical approval statement

The study was conducted with the approval of the Ethics Committee of Beijing Youan Hospital, Capital Medical University and the Ethics Committee of Beijing Tongren Hospital, Capital Medical University.

---

**Table S1:** Demography, clinical and laboratory parameters of patients with COVID-19.

|  | All patients<br>(n=87) | Non-severe<br>(n=67) | Severe<br>(n=20) | P value |
| --- | --- | --- | --- | --- |
| **Age—yrs** | 50 (38-64) | 47 (36-60) | 66 (49-76) | **.000†** |
| **Male sex—n (%)** | 38 (43.7) | 28 (41.8) | 10 (50.0) | .516# |
| **Symptoms** |  |  |  |  |
| **Fever—n (%)** | 64 (73.6) | 50 (74.6) | 14 (70.0) | .681# |
| **Cough—n (%)** | 53 (60.9) | 39 (58.2) | 14 (70.0) | .343# |
| **Sputum—n (%)** | 21 (24.1) | 14 (20.9) | 7 (35.0) | .116! |
| **Dyspnea—n (%)** | 6 (6.9) | 2 (3.0) | 4 (20.0) | **.014!** |
| **Fatigue—n (%)** | 33 (37.9) | 28 (41.8) | 5 (25.0) | .351# |
| **Dysorexia—n (%)** | 10 (11.5) | 6 (9.0) | 4 (20.0) | .111! |
| **Concomitances** |  |  |  |  |
| **Diabetes—n (%)** | 7 (8.0) | 5 (7.5) | 2 (10.0) | .658! |
| **Hypertension—n (%)** | 18 (20.7) | 11 (16.4) | 7 (35.0) | .112! |
| **Routine blood tests** |  |  |  |  |
| **Haemoglobin—g/l** | 138 (125-145) | 139 (128-145) | 133 (121-148) | .366† |
| **Platelet count—$\times 10^9/l$** | 190 (157-233) | 192 (162-233) | 158 (137-233) | .144† |
| **White blood cell count—$\times 10^9/l$** | 4.1 (3.5-5.6) | 4.1 (3.5-5.3) | 5.6 (3.5-7.2) | .142† |
| **Neutrophil percent—%** | 64.3 (51.5-72.1) | 61.7 (50.5-67.9) | 75.7 (66.8-84.1) | **.000†** |
| **Lymphocyte percent—%** | 25.7 (18.8-35.4) | 28.7 (21.7-36.7) | 16.8 (9.4-23.6) | **.000†** |
| **Biochemical parameters** |  |  |  |  |
| **C-reactive protein—mg/l** | 17.4 (3.3-43.5) | 11.3 (2.4-24.7) | 52.7 (33.3-118.1) | **.000†** |
| **Alanine aminotransferase—U/l** | 28 (21-46) | 28 (21-46) | 28 (23-51) | .634† |
| **Aspartate aminotransferase—U/l** | 30 (22-42) | 28 (22-38) | 41 (30-66) | **.005†** |
| **Albumin—g/l** | 36.7 (33.0-39.8) | 37.7 (34.8-40.0) | 33.0 (29.5-34.4) | **.000†** |
| **Creatine kinase—U/l** | 78 (46-123) | 70 (46-118) | 89 (49-264) | .181† |
| **Creatine kinase—MB—ng/ml** | 0.29 (0.14-0.71) | 0.27 (0.11-0.48) | 0.64 (0.26-1.40) | **.003†** |
| **Myoglobin—ng/ml** | 46.5 (30.3-72.5) | 36.0 (29.5-57.0) | 83.0 (52.0-182.0) | **.000†** |
| **Creatinine—$\mu\text{mol/l}$** | 64.5 (55.0-77.8) | 64.0 (54.5-75.0) | 66.0 (55.0-88.0) | .366† |

Data are median (IQR) or n (%). P values were calculated by Mann-Whitney U test (†), $\chi^2$ test (#), or Fisher's exact test(!), as appropriate for group comparison analyses.

**Table S2:** Demography and laboratory parameters of heathy volunteers.

|  | **Healthy**<br>**(n=125)** |
| --- | --- |
| **Age—yrs** | 49 (38-55) |
| **Male sex—n (%)** | 57 (45.6) |
| **Biochemical parameters** |  |
| **Hemoglobin A1c (%)** | 5.3 (5.1-5.5) |
| **Alanine aminotransferase—U/l** | 16 (12-23) |
| **Aspartate aminotransferase—U/l** | 19 (16-23) |
| **Albumin—g/l** | 45.1 (43.5- 47.4) |
| **Creatine kinase—U/l** | 93 (74-128) |
| **Creatinine—μmol/l** | 68.4 (59.3-78.6) |
| **Total cholesterol (mmol/l)** | 4.84 (4.11-5.45) |
| **HDL cholesterol (mmol/l)** | 1.27 (1.14-1.54) |
| **LDL cholesterol (mmol/l)** | 2.97 (2.38-3.44) |
| **Triglycerides (mmol/l)** | 1.17 (0.81-1.76) |

Data are median (IQR) or n (%).

**Table S3:** Circulating parameters correlated with SARS-CoV-2 in patients with COVID-19.

|  | **All patients**<br>**(n=87)** | **Non-severe**<br>**(n=67)** | **Severe**<br>**(n=20)** | ***P* value** |
| --- | --- | --- | --- | --- |
| **ACE2—ng/ml** | 9.6 (7.8-11.5) | 9.4 (7.4-11.7) | 10.3 (8.5-11.5) | .292† |
| **Ang (1-7) —ng/ml** | 0.54 (0.34-0.81) | 0.58 (0.37-0.91) | 0.45 (0.24-0.56) | **.020**† |
| **CTSL—ng/ml** | 1.71 (1.00-3.44) | 1.44 (0.88-2.55) | 4.03 (1.87-7.83) | **.000**† |
| **CTSB—ng/ml** | 0.52 (0.36-0.73) | 0.56 (0.38-0.74) | 0.39 (0.27-0.58) | **.020**† |

2 Data are median (IQR). P values were calculated by Mann-Whitney U test (†) for group comparison analyses.

**Table S4.** Nonparametric correlations of parameters correlated with SARS-CoV-2 infection and severity of the disease.

|  | Severity | CTSL | CTSB | ACE2 | Ang (1-7) | Age | Gender | Diabetes | HBP |
| --- | --- | --- | --- | --- | --- | --- | --- | --- | --- |
| Severity | 1.00 |  |  |  |  |  |  |  |  |
| CTSL | **0.44 (0.000)** | 1.00 |  |  |  |  |  |  |  |
| CTSB | **-0.24 (0.030)** | -0.07 (0.564) | 1.00 |  |  |  |  |  |  |
| ACE2 | -0.11 (0.315) | 0.15 (0.173) | -0.07 (0.543) | 1.00 |  |  |  |  |  |
| Ang (1-7) | -0.21 (0.064) | **-0.26 (0.018)** | 0.07 (0.568) | 0.05 (0.655) | 1.00 |  |  |  |  |
| Age | **0.37 (0.000)** | **0.62 (0.000)** | **-0.22 (0.045)** | **0.24 (0.029)** | **-0.46 (0.000)** | 1.00 |  |  |  |
| Gender | 0.04 (0.718) | 0.02 (0.886) | 0.12 (0.304) | 0.11 (0.351) | 0.02 (0.829) | 0.05 (0.640) | 1.00 |  |  |
| Diabetes | 0.05 (0.658) | 0.20 (0.079) | -0.22 (0.053) | -0.02 (0.871) | -0.04 (0.704) | 0.14 (0.205) | -0.01 (0.964) | 1.00 |  |
| HBP | 0.14 (0.189) | **0.37 (0.001)** | -0.15 (0.188) | 0.06 (0.620) | -0.21 (0.065) | **0.46 (0.000)** | 0.01 (0.942) | **0.37 (0.000)** | 1.00 |

Data are correlation coefficient (p value). Spearman's rho test (2-tailed).

**Figure S1:**
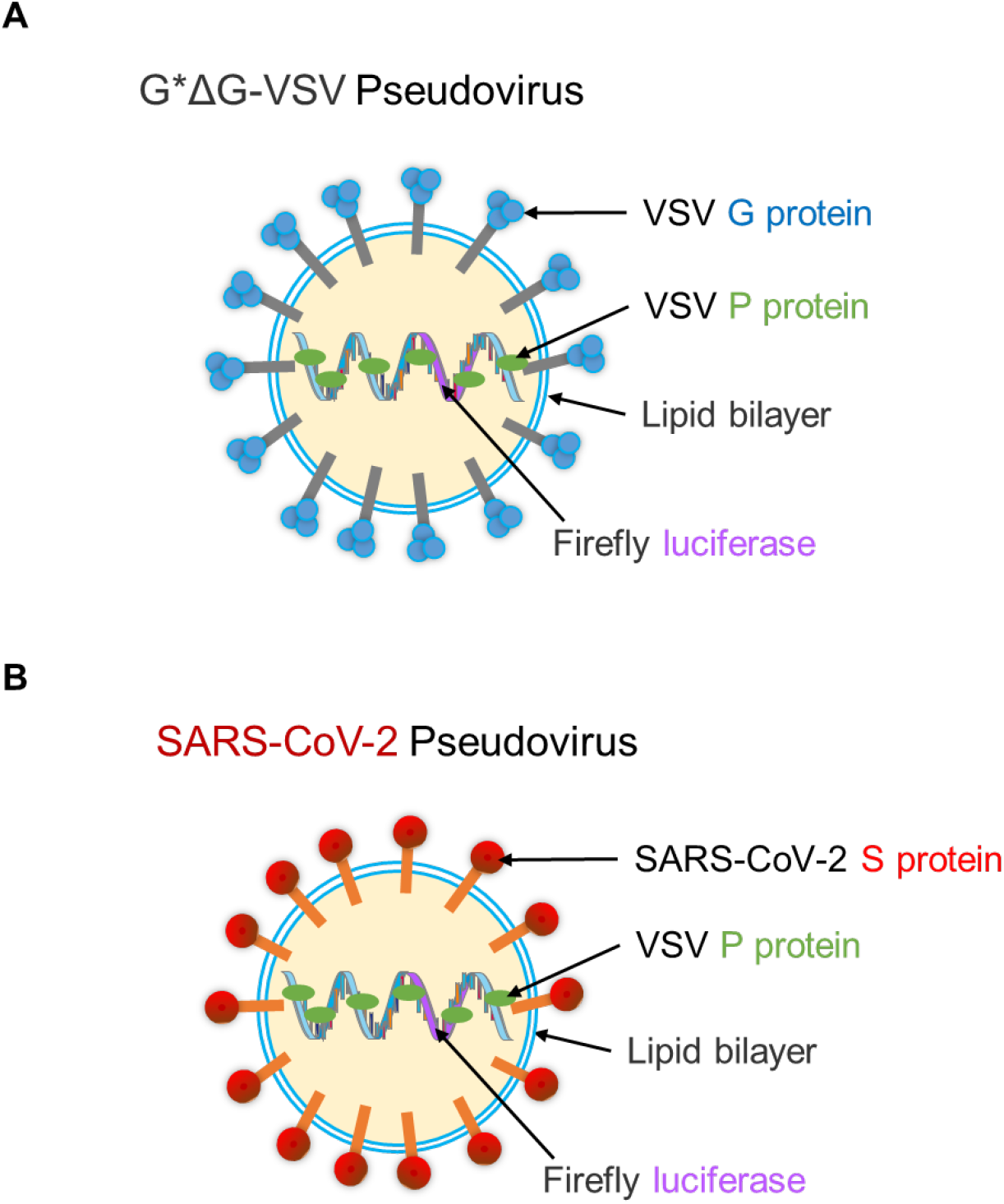
Schematic diagram of the pseudovirus structure. **(A)** The G*ΔG-VSV pseudovirus was generate using a recombinant VSV in which the glycoprotein (VSV-G) gene was deleted and replaced with genes encoding firefly luciferase. When VSV-G is expressed transiently in cells infected with these recombinants, VSV pseudotype particles are produced. **(B)** As VSV virions have no specific for selecting the type of membrane protein that can be incorporated into the viral envelope and VSV particles can bud in the absence of G protein. The SARS-CoV-2 pseudovirus was generated by incorporation of the SARS-CoV-2 S protein into the recombinant VSV stated above.

**Figure S2:**
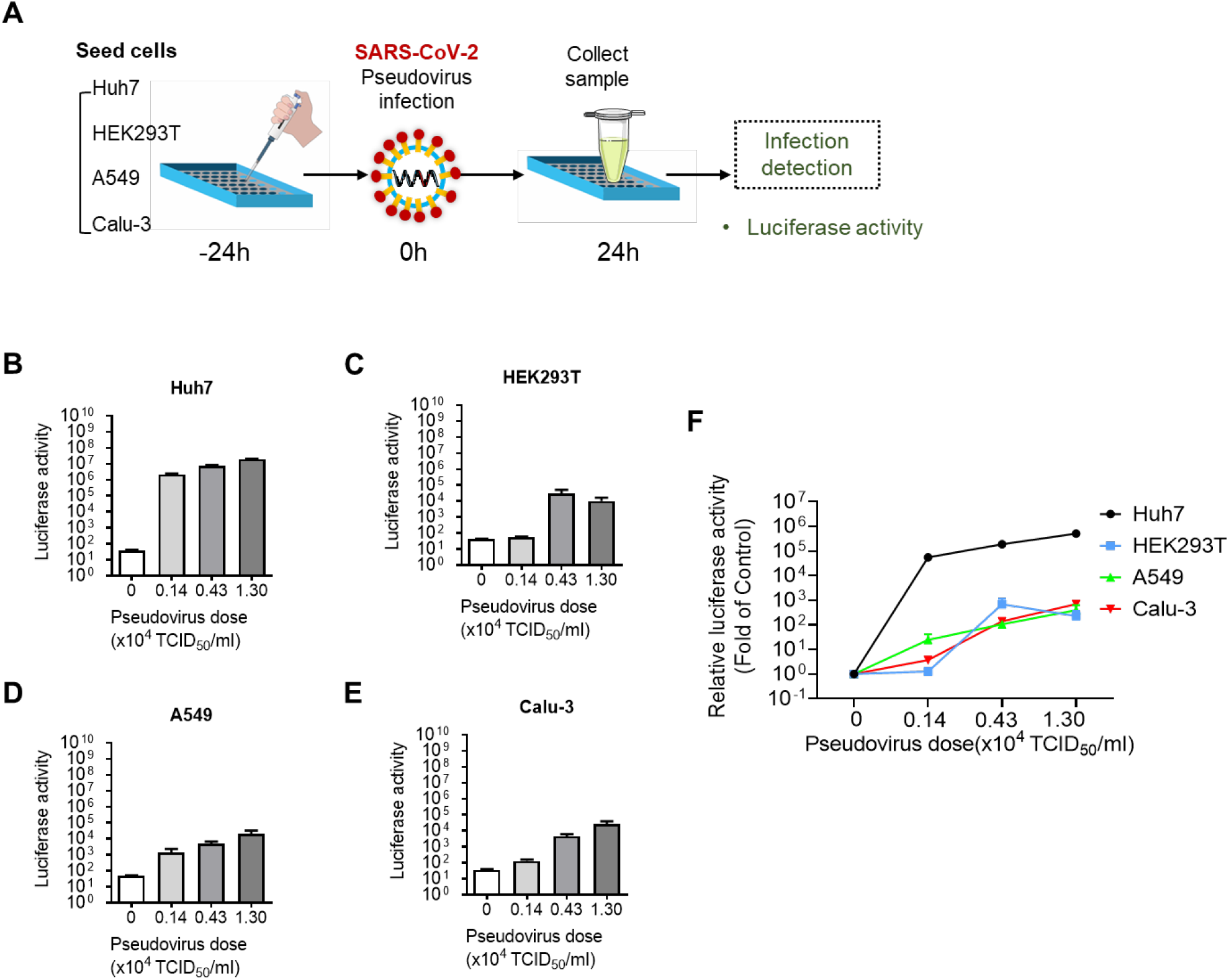
SARS-CoV-2 pseudovirus infection in different human cell lines. **(A)** Schematic of the cell line selection assay setup. Huh7, HEK293T, A549 and Calu-3 cells were infected with different doses of SARS-CoV-2 pseudovirus (from 0.14 to 1.30×10^4^ TCID50/ml) for 24 h. Cells not infected with pseudovirus were used as control cells. **(B-E)** Pseudovirus infection as evaluated by a luciferase assay and shown as absolute luciferase activity values for Huh7 (B), HEK293T (C), A549 (D) and Calu-3 (E) cells. **(F)** Pseudovirus infection as indicated by the relative luciferase activity values in Huh7, HEK293T, A549 and Calu-3 cells. Luciferase activity values were normalized to those in the corresponding control cells. n=4. The data are expressed as the mean ± s.e.m. values.

## STAR*METHODS

### KEY RESOURCES TABLE

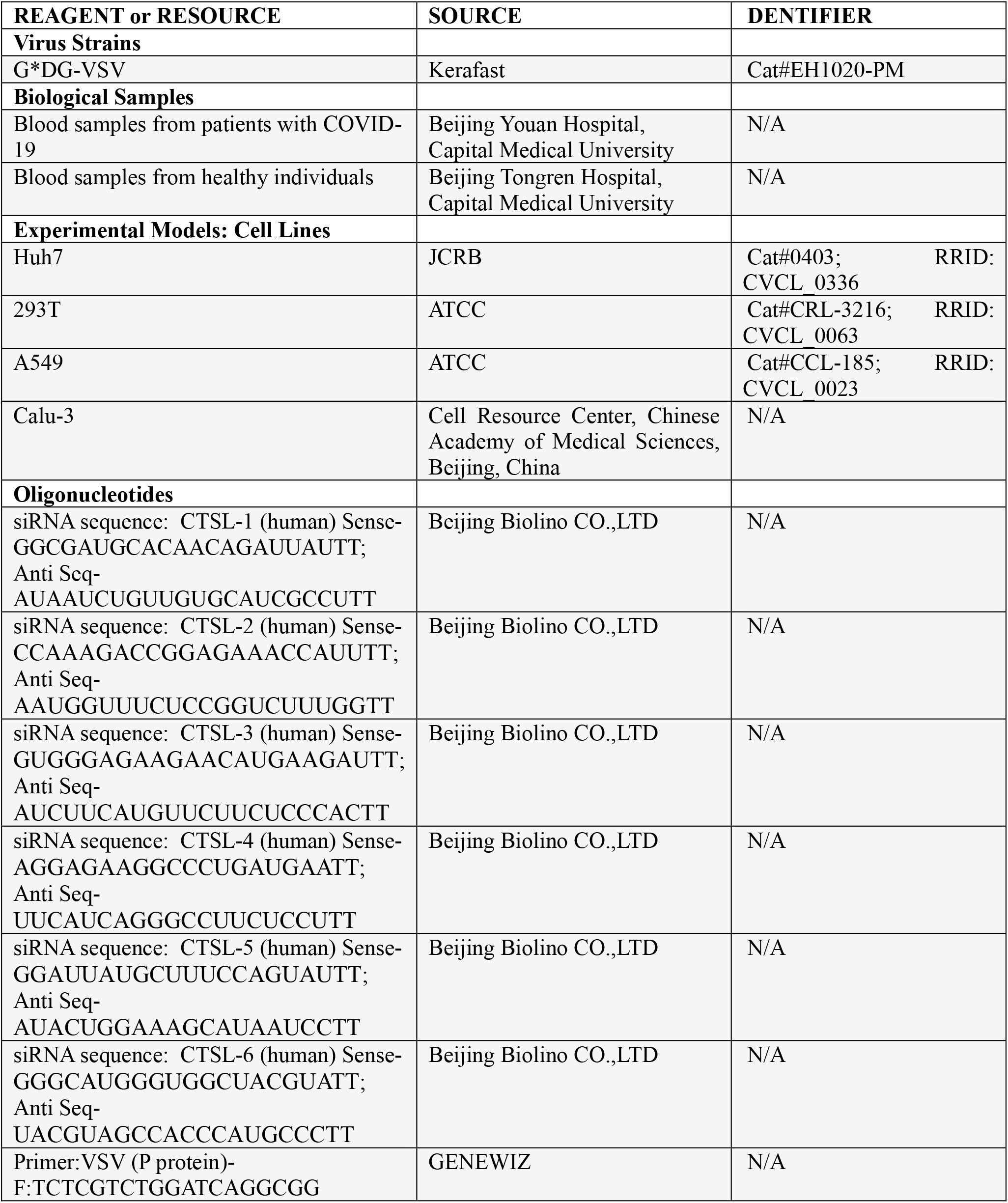

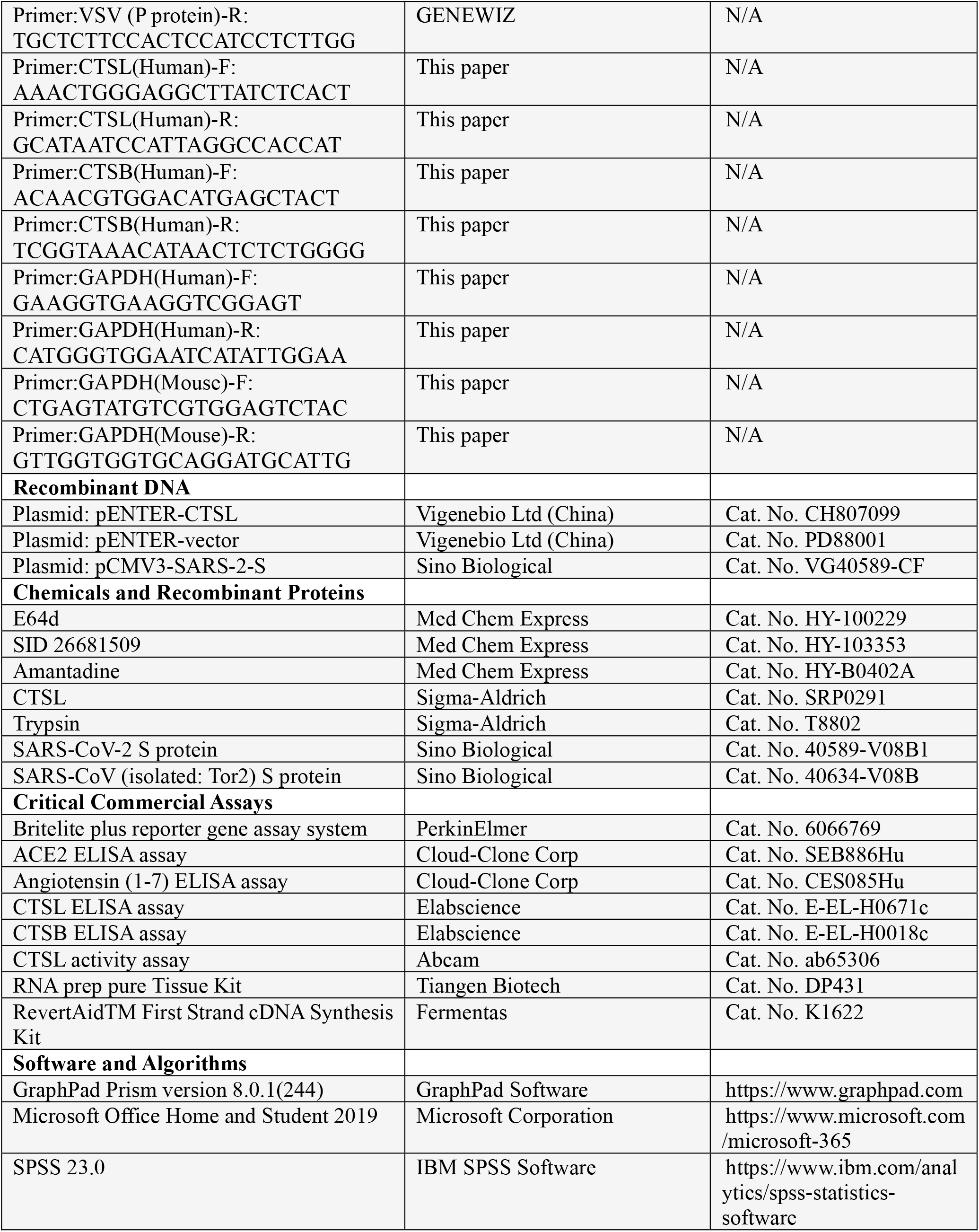

## RESOURCE AVAILABILITY

### Lead Contact

The data that support the findings of this study are available from the Leading Contact, Dr. Jin-Kui Yang.

### Materials Availability

All the unique reagents generated in this study are available from the Lead Contact with a completed Materials Transfer Agreement.

### Data and Code Availability

This study did not generate any unique datasets or code.

## EXPERIMENTAL MODELS AND SUBJECT DETAILS

### Participants and clinical samples

Patients diagnosed with COVID-19 and hospitalized in Beijing Youan Hospital, Capital Medical University from January 21 to April 30, 2020 were enrolled in this study. All enrolled patients were confirmed positive for SARS-CoV-2 nucleic acid by real-time polymerase chain reaction (RT-PCR). The RT-PCR assay was conducted as per the protocol established by the World Health Organization (WHO). Diagnosis and clinical classification criteria and treatment plan (version 7.0) of COVID-19 was launched by the National Health Committee of China (http://www.nhc.gov.cn/). The clinical classification of severity is as follows: (1) *Mild*, having only mild symptoms, imaging shows no pneumonia. (2) *Moderate*, with fever, respiratory tract symptoms, and imaging shows pneumonia. (3) *Severe*, meet any of the following signs: a) respiratory distress, respiratory rate ≥ 30 beats / min; b) in the resting state, finger oxygen saturation ≤ 93%) arterial blood oxygen partial pressure (PaO_2_/oxygen concentration (FiO_2_) ≤ 300mmHg (1mmHg = 0.133kPa). (4) *Critical*, one of the following conditions: a) respiratory failure occurs and requires mechanical ventilation; b) Shock occurs; c) ICU admission is required for combined organ failure. Patients with COVID-19 experienced a mean of 14 days of hospitalization (Day 14) and were followed up on the 14^th^ day (Day 28) and 28^th^ day (Day 42) after discharge from hospital. Blood samples were collected shortly after the admission to the hospital (for some patients who were transferred from other hospitals, their blood samples were collected shortly after admission at Beijing Youan hospital) and on Day 28 and Day 42. Demographic, clinical, and laboratory data were extracted from the electronic hospital information system using a standardized form.

A total of 125 sex- and age-matched healthy volunteers were recruited in Beijing Tongren Hospital, Capital Medical University. The including criteria are as followed: 1) Age 18-70y; 2) No underlying diseases; 3) No long-term use of any medication; 4) Willing to participate in the study. Blood samples were collected after an overnight fasting for the determination of biochemical parameters, CTSL and cathepsin B CTSB concentrations. All biochemical measurements have been participated in the Chinese Ministry of Health Quality Assessment Program. This study was conducted with the approval of the Ethics Committee of Beijing Youan Hospital, Capital Medical University and the Ethics Commission waived the requirement for informed consent.

### Cell lines and reagents

The human hepatoma cell line Huh7, human lung adenocarcinoma A549 cells and human HEK293T(293T) cells were maintained in high glucose Dulbecco’s modified Eagle’s medium (DMEM) (Sigma-Aldrich, St. Louis, MO, USA) supplemented with 10% fetal bovine serum (FBS, Gibco, Carlsbad, CA), 100 units/ml penicillin and 100 mg/ml streptomycin (Thermo Fisher Scientific). The human Calu-3 lung adenocarcinoma cell line were cultured in minimum essential medium (Eagle) with 2 mM L-glutamine and Earle’s BSS adjusted to contain 1.5g/l sodium bicarbonate, 0.1 mM non-essential amino acids and 1.0 mM sodium pyruvate and 10% FBS. All the cells were maintained at 37°C in a humidified atmosphere containing 95% air and 5% CO_2_.

### Experimental Mice

The study used 4-5-week-old human ACE2 transgenic mice (weight 13-17g), a mouse model expressing human ACE2 (hACE2) generated by using CRISPR/Cas9 knockin technology as previous reported (Sun et al., 2020). All animal protocols were approved by the Ethical Review Committee at the Institute of Zoology, Capital Medical University, China.

## METHOD DETAILS

### Serum biomarkers of patients with COVID-19

Plasma samples of patients with COVID-19 collected at admission (d0), 14th day after discharge (d28) and 28th day after discharge (d42) were collected and stored at -80°C within 2h. Angiotensin (1-7), ACE2, CTSL and CTSB were analyzed using commercially available enzyme-linked immunosorbent assays (ELISA) following the manufacturer’s instructions. All samples were detected without virus inactivation to retain the original results in a P2+ biosafety laboratory.

### Pseudovirus

The SARS-CoV-2, SARS, RVF and VSV pseudovirus were generated with the incorporation of SARS-CoV-2 S protein, SARS-CoV-1 S protein, RVF G protein, and VSV G protein into VSV-based pseudovirus system, respectively. For this VSV-based pseudovirus system, the backbone was provided by VSV G pseudotyped virus (G*ΔG-VSV) that packages expression cassettes for firefly luciferase instead of VSV-G in the VSV genome (Nie et al., 2020). Therefore, the luciferase activity and the mRNA level of VSV phosphoprotein (VSV-P) were used for indicators of pseudovirus infection.

### Luciferase assay

The activities of firefly luciferases were measured on cell lysates using luciferase substrate following the manufacturer’s instructions. Briefly, for 96-well plates, the culture supernatant was aspirated gently to leave 100μl in each well; then, 100μl of luciferase substrate was added to each well. Two minutes after incubation at 37°C, 150μl of lysate was aspirated to a clean 1.5 ml sterile EP tube to measuring the firefly luciferase activity for each well rapidly using a luminometer (Turner BioSystems) as described previously(Yang et al., 2017).

### Cell line selection and clinical data verification *in vitro*

Huh7, 293T, A549 and Calu-3 cells were plated in 48-well plates respectively and infected with different dose of SARS-CoV-2 pseudovirus (starting from 0 to 1.3×10^4^TCID_50_/ml). The cells were cultured for another 24 hours before luciferase activity analysis. Cells without the addition of pseudovirus as the cell control. The most susceptible cell line was selected for subsequent experiments. To verify the clinical data, Huh7 cells were plated in 48-well plates, and allowed to adhere until the cells are about 70% confluent, followed by infecting with different dose of SARS-CoV-2 pseudovirus (starting from 0 to 1.3×10^4^TCID_50_/ml). After 24 hours incubation, the cells were lysed for analysis the firefly luciferase activity, VSV-P mRNA and the detection of CTSL and CTSB by ELISA assays.

### Cleavage of SARS-CoV-2 S protein by CTSL

Purified ectodomain of SARS-CoV-2 S protein (NCBI reference sequence YP_009724390.1, residues 16-1213) and ectodomain of SARS-CoV (isolated: Tor2) S protein (NCBI reference sequence NP_828851.1, residues 1-1195) were expressed in baculovirus-insect cells. One microgram of each protein was incubated with 2 or 10 μg/ml CTSL in assay buffer (400 mM sodium acetate, pH 5.5, with 4 mM EDTA and 8 mM DTT) for 1h at 37°C. CTSL was preactivated in 30°C for one minute before use. Where indicated, 20 μM E64d or 20 μM SID26681509 was added in the reaction system with 0.5 μg SARS-CoV-2 S protein and 2 μg/ml CTSL. The proteins were then subjected to sodium dodecyl sulfatepolyacrylamide gel electrophoresis (SDS-PAGE) and analyzed by silver stain.

## Syncytium formation assay

Huh7 cells were seeded in 24-well plates and transfected with SARS-CoV-2 S protein expression plasmids (NCBI reference sequence YP_009724390.1, QHD43416.1) (0.65 μg/well) using Lipofectamine 3000 reagent. The transfection solutions were changed to standard culture medium 6 h post-transfection and cells incubated for additional 12 h. Next, cells were treated in the absence (PBS, pH=7.4) or presence of 2 μg/ml trypsin (Sigma-Aldrich) (in PBS, pH=7.4), or in the absence (PBS, pH=5.8) or presence of 2 or 4 μg/ml CTSL (Sigma-Aldrich) (in PBS, pH=5.8) for 20 min at 37°C. Then, the solutions were changed to standard culture medium and the cells further incubated for 16 h. The pictures were captured under bright-field microscopy (Olympus) and analyzed the formation of syncytia by counting the nuclei in syncytia in five random microscopic fields.

### CTSL knockdown by siRNA and overexpression by plasmid *in vitro*

For CTSL knockdown, Huh7 cells were plated in 48-well plates, and transfected with 50 nM or 100 nM siRNAs against homo CTSL mRNA (si-CTSL) or 50 nM negative control siRNA (scramble) using Lipofectamine 3000 reagent. For CTSL overexpression, Huh7 cells were plated in 48-well plates, and transfected with 0.2 μg or 0.4 μg human CTSL expression plasmid (pENTER-CTSL, pCTSL) or 0.2 μg control plasmid (pENTER-vector, Con). 24 hours post transfection, the cells were lysed for analysis the CTSL and CTSB mRNA level to evaluate the efficiency of si-CTSL and pCTSL. To evaluate the effect of CTSL on SARS-CoV-2 entry, Huh7 cells were plated in 48-well plates, and transfected with si-CTSL or pCTSL under the same conditions stated above. 24 hours post transfection, the medium was replaced with fresh medium. Then the cells were infected with SARS-CoV-2 pseudovirus (1.3×10^4^TCID_50_/ml) and cultured for another 24 hours before firefly luciferase activity and VSV-P mRNA analysis.

### Effect of drug treatment on SARS-CoV-2 entry *in vitro* and cell viability assay

The anti-SARS-CoV-2 activity of SID26681509, E64d and amantadine were performed in 96-well plates by quantification of the firefly luciferase activity. Huh7 cells were pretreated with different concentrations of drug or the equivalent amount of solvent for 1 hour and then infected with SARS-CoV-2 pseudovirus (1.3×10^4^TCID_50_/ml) in a 5% CO_2_ environment at 37°C for 24 hours before firefly luciferase activity analysis. In detail, the concentrations of different drugs as follow: SID26681509(0.2μM, 2μM, 4μM, 20μM, 40μM and 100μM), E64d (0.14μM, 0.42μM, 1.23μM, 3.7μM, 11.1μM and 33.3μM), and amantadine (1.56μM, 6.25μM, 25μM,100μM, 400μM and 1600μM). The effects of SID26681509, E64d and amantadine on cell viability were measured by MTT assay. Huh7 cells were seeded into a 96-well plate at a cell density of 0.5×10^4^ per well and allowed to adhere until the cells are about 70% confluent, followed by treatment with different concentrations of drugs or the equivalent amount of solvent for 24 hours. The concentrations of different drugs were detailed above. Cells without any treatments as the blank control. After treatments, MTT was added into the culture medium to the final concentration of 0.5mg/ml, and then the cells were incubated for 4 hours at 37°C in an incubator. After removing the culture medium, the cells were lysed by gently rotating in 200μl DMSO for 10 minutes in darkness at room temperature. The absorbance at 570nm was measured using an automatic plate reader. The average absorbance reflected cell viability with the data normalized to the blank control group. Experiments were done in quintuplicates and repeated at least three times.

### Animal experiments

The anti-SARS-CoV-2 activity of E64d and amantadine were performed in human ACE2 transgenic mice by bioluminescent imaging (BLI) assay as before (Zhang et al., 2017). In brief, 4-5-week-old human ACE2 transgenic mice were treated with E64d (12.5mg/kg body weight) or amantadine (50mg/kg body weight) or the equivalent amount of solvent once a day via the intraperitoneal (IP) route 2 days prior to virus inoculation. Then mice were injected with 1.5×10^6^ TCID_50_ SARS-CoV-2 pseudovirus per mouse via tail vein injection. Mice pre-treated with drug solvent but without pseudovirus inoculation served as the healthy control group. Bioluminescence was measured one day post-infection and visualized in pseudocolor. Finally, mice were sacrificed for experimental analysis immediately after bioluminescence measurement.

### RNA extraction and Real-time PCR

Total RNA was extracted from cultured cells or mouse livers and the reverse transcription was performed. The real-time qPCR was then performed on the LightCycler® 96 Real-Time PCR System (Roche) using SYBR Green I Master Mix reagent with the primers and using GAPDH as the house-keeping gene.

## QUANTIFICATION AND STATISTICAL ANALYSIS

Clinical data were expressed as median (interquartile range (IQR)) or percentage, as appropriate. Comparison of continuous data between groups were determined using Mann-Whitney *U* test. Chi-square (χ^2^) test or Fisher’s exact tests were used for categorical variables as appropriate. To explore the risk factors associated with severity, univariate and multivariate logistic regression models were used. Spearman’s rho test (2-tailed) were used to analyze nonparametric correlations of parameters correlated with SARS-CoV-2 infection and severity of the disease. SPSS for Windows 23.0 and Graphpad prism 7.0 software were used for statistical analysis, with statistical significance set at 2-sided P<0.05.

